# Gut microbiome predicts atopic diseases in an infant cohort with reduced bacterial exposure due to social distancing

**DOI:** 10.1101/2023.03.22.23287583

**Authors:** Katri Korpela, Sadhbh Hurley, Sinead Ahearn Ford, Ruth Franklin, Susan Byrne, Nonhlanhla Lunjani, Brian Forde, Ujjwal Neogi, Carina Venter, Jens Walter, Jonathan Hourihane, Liam O’Mahony

## Abstract

Several hypotheses link altered microbial exposure in affluent societies to increased prevalence of allergies, but none have been experimentally tested in humans. Here we capitalize on the opportunity to study a cohort of infants (CORAL) raised during COVID-19 associated social distancing measures to test the interactions between bacterial exposure and fecal microbiome composition with atopic outcomes. We show that fecal *Clostridia* levels were significantly lower in CORAL infants and correlated with a microbial exposure index. Microbiota composition was the most significant component of regression models predicting risk of atopic dermatitis (AUC 0.86) or food allergen sensitization (AUC 0.98) and mediated the effects of multiple environment factors on disease risk. Although diet had a larger effect on microbiota composition than environmental factors linked to dispersal, most effects were mediated through the microbiota. This study provides critical information to refine existing hypothesis on the importance of the gut microbiota to immune development.

## Introduction

Human mucosal surfaces and body cavities harbor diverse communities of commensal microbes that play essential roles in regulation of host metabolic responses, epithelial barrier function, immune education, and immune regulation (1–6). Humans evolved in an environmental and social context that enabled reliable transmission and dispersal of human adapted symbionts, accompanied by diets rich in substrates that provide nutritional support for the gut microbiota (7,8). Hallmarks of these evolutionary relationships can be observed in the microbiomes of non-industrialized human populations and non-human primates, whose microbiomes are shaped by dispersal and social behavior (9–11). Given that the gut microbiota provides essential, and evolution routed benefits to human development, alterations in microbial transmission through lifestyle factors linked to industrialization and western dietary habits provide a possible mechanism for the rise in chronic, non-communicable diseases in affluent societies.

There is an “immunological window of opportunity” early in life when the immune system is particularly responsive to environmental exposures (including infections, nutrition, and microbiome) that help establish the thresholds, patterns of reactivity, and functional trajectories that can have long term consequences including altered risk of immune mediated diseases (12–15). The immune response to commensal microbes is not simply a form of host defense but represents an intimate and sophisticated bidirectional dialogue that ensures a stable microenvironment is maintained with important symbiotic physiological effects on the host. These microbe-host interactions promote appropriate immune responses that effectively defend against infection, with limited collateral damage to host tissue, and without any subsequent aberrant inflammatory or allergic reactions (16–20).

Epidemiological factors linking immune-mediated diseases with living environment, lifestyle and social interactions (e.g. household size, absence of older siblings, antibiotics, mode of delivery, breast feeding rates, rural versus urban living, dietary intake, etc.) are also known to impact early-life gut microbiota assembly (21–24). These environmental factors alter ecological processes that shape community assembly, such as dispersal (household size, siblings, c-sections) and selection (antibiotics, breast feeding, diet). Metacommunity theory predicts that the composition and dynamics within any local community (e.g. within a human individual) are governed both by processes that occur within the community (dietary resources, antibotics) and by the process of dispersal (siblings, hygiene, family size) which links communities together (25–27). These theoretical frameworks led us to hypothesize that allergies, and the increase in allergy with industrialization, arise from modern lifestyle-factors disrupting human gut metacommunity composition and function.

The importance of environmental factors that influence metacommunity dynamics through dispersal (family size, siblings, social interactions) can usually only be examined in observational studies. However, the enforced social distancing measures that were employed during sustained periods of mass lockdowns to prevent spread of the SARS-CoV-2 virus provided a real-world unique opportunity to experimentally test the metacommunity hypothesis of atopic disease. The CORAL (Impact of **<u>COR</u>**ona Virus Pandemic on **<u>Al</u>**lergic and Autoimmune Dysregulation in Infants Born During Lockdown) study examined effects of the social isolation restrictions imposed during the early stages of the COVID-19 pandemic on infants that spent most of their early life in social isolation. Compared with a historical Irish cohort, infants in the CORAL cohort had higher rates of atopic dermatitis (25% versus 17%) and food allergen sensitization (7% versus 4%) at 12 months of age (28). Furthermore, some deficits in social communication were evident at their 12-month assessment (29). Here we extend this work, hypothesizing that environmental factors that alter metacommunity processes affected microbiome composition, and that these interactions might mediate allergic outcomes, which are some of the earliest manifestations of inappropriate immune reactivity. To test this hypothesis, we performed 16S rRNA tag sequencing on fecal DNA and applied an ecological modeling approach to determine the relative importance of the environmental drivers of bacterial community assembly. We then determined to what degree interactions between bacterial taxa and dietary factors could predict levels of atopic dermatitis and food allergen sensitization in this population.

## Results

### CORAL infant cohort characteristics

CORAL demographic details have been previously published (28, 30). In summary 55% were male, 96% Caucasian, 46% first born infants, 38% second born or later infants, 65% were born by spontaneous vaginal delivery. Despite multiple waves of COVID-19 in the community during this time, social isolation was effective in minimizing exposure to SARS-CoV-2 as only 3% of infants had a detectable antibody response to the virus at 12 months of age. In addition, only 17% of infants required antibiotics by 12 months (compared to 80% in a pre-pandemic UK birth cohort) further indicating reduced exposure to infectious agents (31).

Families of infants in the CORAL cohort had a median of one social contact outside the home at birth, increasing to four when the infants reached 6 months of age. The median number of people who had kissed the infant in the first 6 months was 3 (including parents). In addition, social isolation restrictions led to 25% of infants not having met a child their own age by their first birthday (32).

### Fecal bacterial community composition

We sequenced fecal samples from all infants for which background questionnaire data was available at 6 (n=351) and 12 months (n=343), using 16S rRNA tag sequencing. Consistent with previous studies, infant microbiota composition was strongly associated with age and birth mode. Age explained 26% of the variation in composition between samples (PERMANOVA, p=0.001) and birth mode explained 3% at 6 months (PERMANOVA, p=0.001) and 1.5% at 12 months (p=0.004) (**Supplementary Fig 1a**). Age was significantly associated with the relative abundances of 29 (33%) of the 89 detected bacterial families (FDR-corrected p<0.05). At the order level, *Bifidobacteriales, Lactobacillales, Enterobacteriales*, and *Veillonellales* decreased from 6 to 12 months of age, while *Verrucomicrobiales, Erysipelotrichales, Clostridiales, Bacteroidales*, and *Coriobacteriales* increased from 6 to 12 months of age (**Supplementary Fig 1b**).

**Fig 1.**
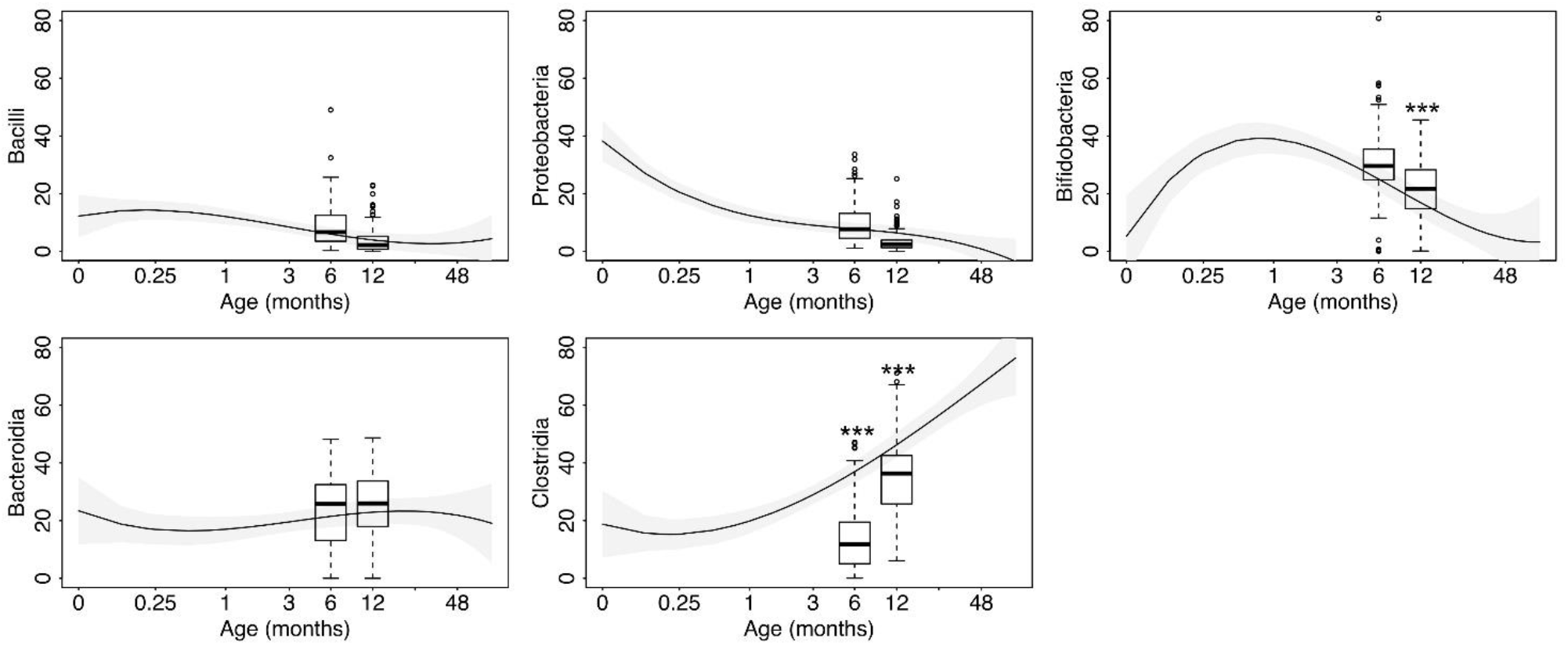
Comparison to the general pattern of microbiota development in other cohorts. The relative abundances of the most abundant bacterial taxa (phylum Proteobacteria, classes *Bacilli, Bacteroidia, Clostridia*, and family *Bifidobacteriacea*). The trend lines are calculated from a third-degree polynomial regression model weighted by the sample size of each study. The boxplots represent the relative abundances in the CORAL cohort. Significance of the differences is marked with asterisks (*** p< 0.001).

Also consistent with previous studies, C-section (CS) delivery or exposure to intrapartum antibiotics during vaginal birth (VAB) were associated with increased relative abundance of the genera *Sarcina* (*Clostridiaceae*), *Flavonifractor* (*Ruminococcaceae*), and *Erysipelatoclostridium* (*Erysipelatoclostridiaceae*) and reduced relative abundance of *Collinsella* (*Coriobacteriaceae*) at 6 months of age. In addition, CS delivery was associated with increased relative abundance of *Enterobacter* (*Enterobacteriaceae*), *Amedibacterium* (*Erysipelotrichaceae*), *Blautia*, and *Roseburia* (*Lachnospiraceae*), and reduced relative abundance of *Bacteroides* (*Bacteroidaceae*) and *Parabacteroides* (*Porphyromonadaceae*) (FDR-corrected p< 0.05). At 12 months, most birth-mode related differences were no longer significant, except for *Bacteroides* that was still significantly reduced in the CS-delivered infants, and *Faecalibacillus* (*Erysipelotrichaceae*) in the VAB group (FDR-corrected p< 0.05).

### CORAL microbiota comparison with historical cohorts

We compared the microbiota composition in the CORAL cohort to the general pattern of microbiota development observed across multiple previously published cohorts, including data from 34 studies, 3825 infants, and 5732 fecal samples (33). Similar time-dependent abundance of *Bacilli, Proteobacteria* and *Bacteroidia* were observed in the CORAL and previous cohorts (**Fig. 1**). However, CORAL infants had significantly lower levels of *Clostridia* at both 6 months and 12 months of age (p<0.001). In addition, the relative abundance of *Bifidobacterium* (class *Actinomycetia*) was elevated at 12 months in comparison to other cohorts (p<0.0001).

We also compared CORAL microbiota data with a recently published Irish infant cohort (INFANTMET) that was studied prior to the pandemic (34, 35). *Clostridia* were also significantly less abundant in the CORAL cohort compared to the INFANTMET cohort at both 6 (p<0.001) and 12 (p=0.02) months of age, while *Bifidobacteria* were more abundant in the CORAL cohort at 12 months of age,(p<0.0001) after adjusting for birth mode, breastfeeding, and antibiotic use. The Clostridia pattern was mainly driven by the reduced relative abundance of the genera *Sarcina* (Clostridiaceae, p<0.0001) and *Eubacterium* (Eubacteriaceae, p<0.0001) at 6 months, *Enterocloster* (*Ruminococcaceae*, p<0.001) and *Hungatella* (Lachnospiraceae, p<0.0001) at both time points, and additionally family *Ruminococcaceae* at 12 months (p<0.001) and the genus *Flavonifractor* (*Ruminococcaceae*, p<0.0001) at 12 months. At 6 months, the CORAL cohort harbored a higher microbial richness than the pre-pandemic cohort (p<0.0001), but the pattern had reversed by 12 months due to the lower rate of increase in richness in CORAL (p<0.0001).

In order to determine if reduced abundance of *Clostridia* could be related to differences in microbial exposures, we examined correlations with a microbial exposure index generated by combining answers to questionnaire data relating to potential sources of microbial exposures (i.e. having a family member classified as an essential worker who was allowed to work outside the home, number of siblings, number of adults in the household, attending daycare outside of home, having pets, and living in a house as compared to an apartment). The relative abundance of *Clostridia* was associated with the exposure index (p=0.039) at 6 months of age **(Fig. 2)**, supporting the hypothesis that reduced exposure to external sources of microbes was linked with the lower-than-expected relative abundance of *Clostridia*. At the genus level, all *Clostridial* genera showed a positive trend with external exposure, which was statistically significant (p<0.05) in 19/38 (50%) of the genera detected at 6 months of age **(Fig. 2)**. At 6 months, the exposure index was also positively associated with microbial richness (p=0.01). The recorded exposures were not significantly associated with Clostridia (p=0.73) or richness (p=0.08) by 12 months of age.

**Fig. 2.**
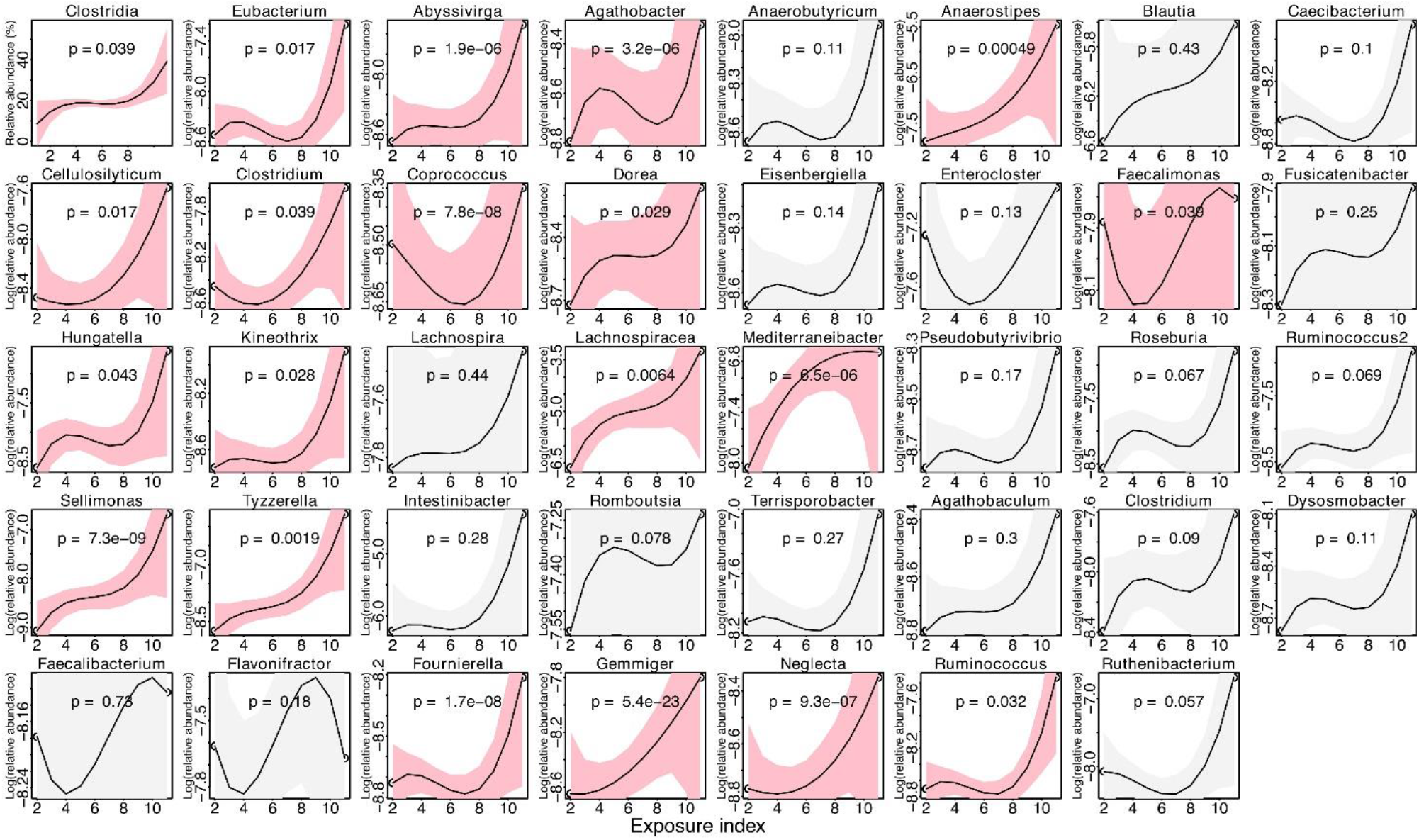
Association of the class Clostridia (first panel), and Clostridial genera with external exposure index. Trendlines, 95% confidence intervals, and p-values are calculated based on a third-degree polynomial regression model. Significant associations are highlighted with pink.

### Relative contribution of environmental factors on microbiome development in the CORAL cohort

We analyzed the contribution of multiple potential contributing factors on the gut microbiota composition at 6 and 12 months, using the envfit function in R package vegan. At both time points, diet was the main determinant of microbiota composition, exceeding even the effect of birth mode (**Fig. 3a**). The measured variables explained 32% and 31% of the variation in microbiota composition at 6 and 12 months, respectively. The impact of medical interventions, primary caesarean delivery, and intrapartum antibiotics were higher at 6 months, while the effects of exposure to other humans and the environmental exposures increased at 12 months (**Fig. 3b and Fig. 3c**).

**Fig.3.**
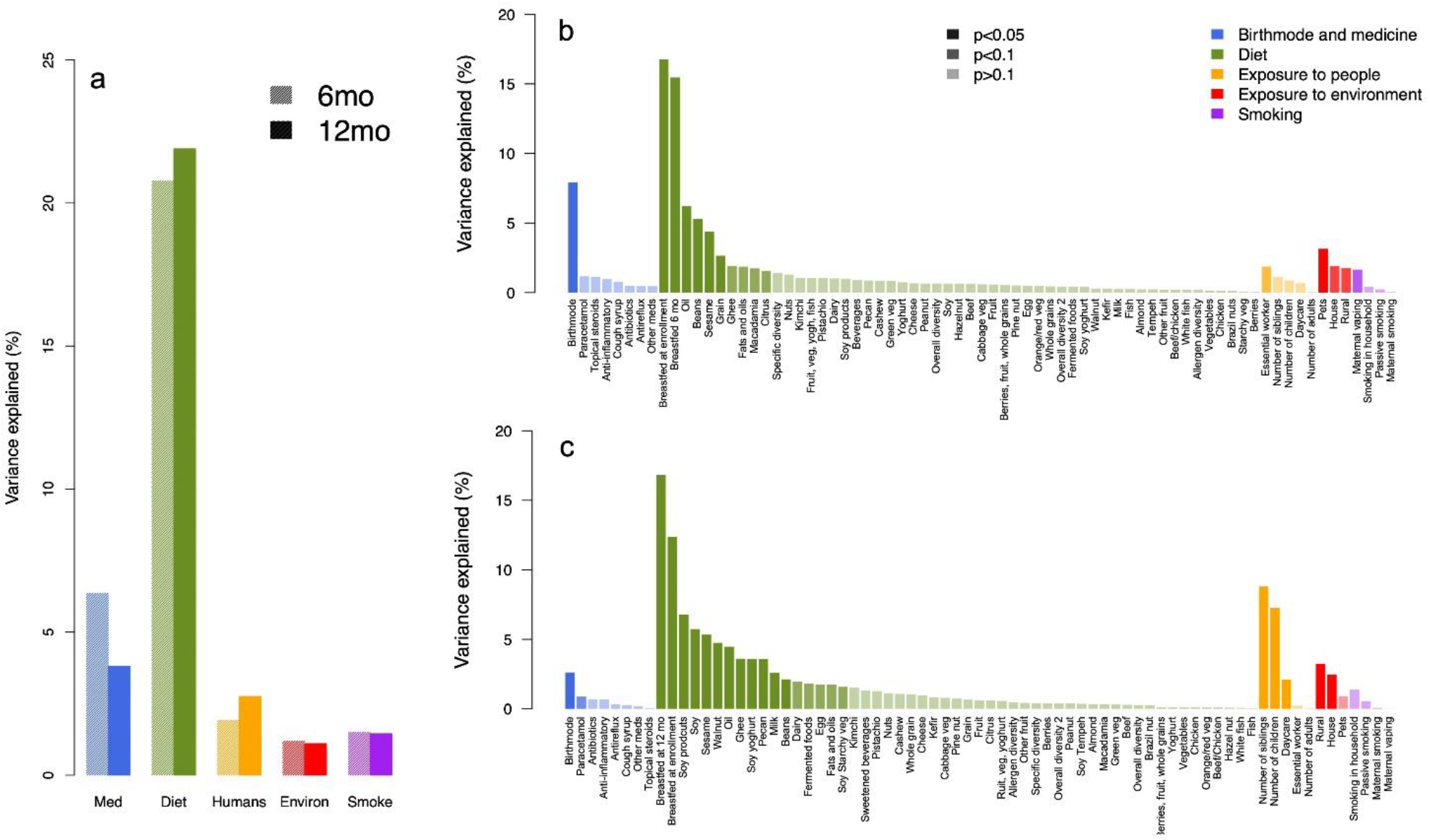
Contribution of different types of exposures on infant gut microbiota with total variance attributable to the different categories (a), and detailed associations at 6 months (b), and 12 months (c).

Of the dietary components, breastfeeding (including exclusive and non-exclusive) had the strongest impact at both time points (12-17% variance explained at both time points). At 6 months, the frequency of consumption of vegetable oil, beans, sesame seeds, and grains had a significant association with microbiota composition (**Fig. 3b, 4a**). At 12 months, additionally soy, walnuts, pecan nuts, and cow’s milk showed a significant contribution (**Fig. 3c, 4b**). At both time points, the effect of oil, beans and nuts was similar in direction to that of breastfeeding and vaginal birth, directing the microbiota towards increased relative abundances of *Bifidobacterium, Lactobacillus, Collinsella*, and *Bacteroides* (**Fig. 4c and Fig. 4d**). Cow’s milk formula consumption, on the other hand, had the opposite effect to breastfeeding and vaginal birth at 12 months, shifting the microbiota away from the *Bifidobacterium/Bacteroides*-dominated composition.

**Fig. 4.**
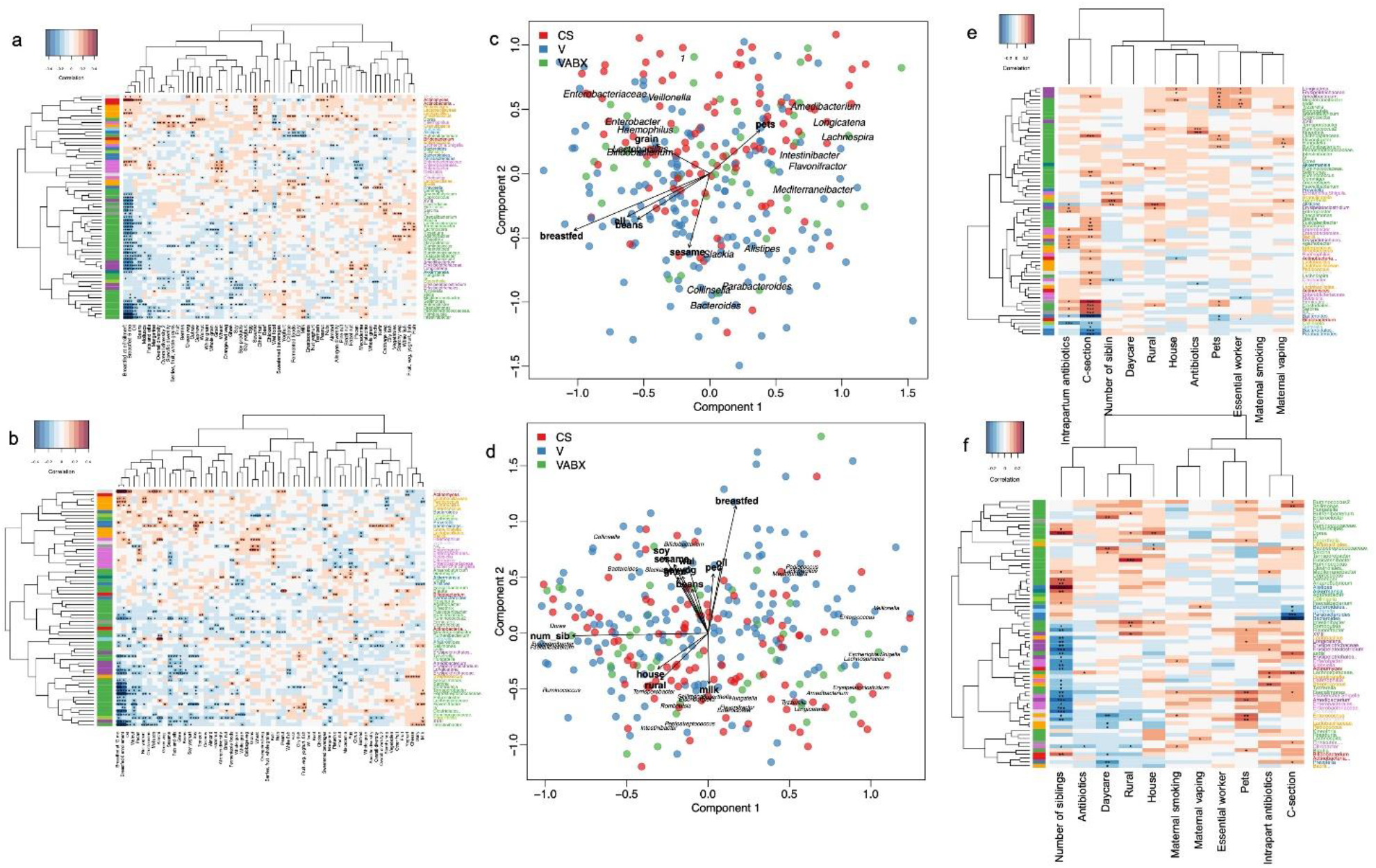
Association between bacterial genera and food categories (a,b), most significant variables (c,d), and external exposures (e,f) at 6 months (a,c,e) and 12 months (b,d,f). Heat maps (a,b,e,f) represent the correlations between log-transformed relative abundances of bacteria, coloured by class, and dietary/exposure variables. Panels (c,d) show the significant associations (p<0.05) of exposure variables with the first two principal coordinates as well as the bacterial genera most strongly associated with the coordinates, with samples coloured by birth mode.

The impact of factors linked to human exposure dramatically increased from 6 to 12 months with effect of the number of siblings increasing from 1% to 9% of the variation. Pets and siblings at 6 and 12 months, respectively, shifted the microbiota from the infant-type *Bacilli-Enterobacteria*-composition towards a more mature composition with increased relative abundance of members of *Ruminococcaceae* and *Lachnospiraceae* (**Fig. 4e**). Pets was the strongest environmental factor at 6 months, while at 12 months, the living environment (rural versus urban, house versus apartment) became more significant, with exposure to a rural living environment shifting the microbiota towards increased relative abundance of the *Clostridial* families *Ruminococcaceae, Lachnospiraceae*, and *Peptostreptococcaceae* (**Fig. 4f**).

### Microbiome mediates the effects of environmental factors on atopic dermatitis and food allergen sensitization

Virtually all the environmental factors identified above as predictors of microbiome variation (diet, breastfeeding, siblings, pets, etc.) have also been described as epidemiological factors that influence atopic dermatitis and food allergen sensitization. To determine the extent to which the microbiota mediates the effects of epidemiological factors, we compared the estimated impact of exposures on disease outcomes with and without adjusting for gut microbiota composition using logistic regression models, adjusting for parental allergic diseases and ethnicity.

The atopic dermatitis prediction model based purely on the environmental factors without gut microbiota data achieved an AUC of 0.82 (95% confidence interval 0.76-0.88). We then selected the most significantly associated microbial genera measured at 6 and 12 months and added those into the model. Adding the gut microbiota data as additional explanatory variables in the model increased the AUC to 0.87 (95% CI 0.82-0.92) (**Supplementary Fig 2**). The gut microbiota composition at both 6 months and 12 months was strongly predictive of atopic dermatitis diagnosis at 12 months (**Fig. 5a**). The atopic dermatitis-associated microbes, apart from *Akkermansia* (*Akkermansiacaeae*) and *Amedibacterium (Erysipelotrichaceae)*, were mostly Clostridia, with all but *Sarcina (Clostridiaceae)* at 6 months having a positive association with atopic dermatitis. In particular *Enterocloster* (*Lachnospiraceae)*, was associated at both timepoints with increased risk of atopic dermatitis. Regarding exposures, atopic dermatitis was positively associated with attending daycare, and smoking in the household, and negatively associated with living in a rural area and with antibiotics use between 6 and 12 months of age. The effect of smoking was mainly direct, rather than microbiota mediated, since including the gut microbiota in the model did not reduce the estimated effect size.

**Fig. 5.**
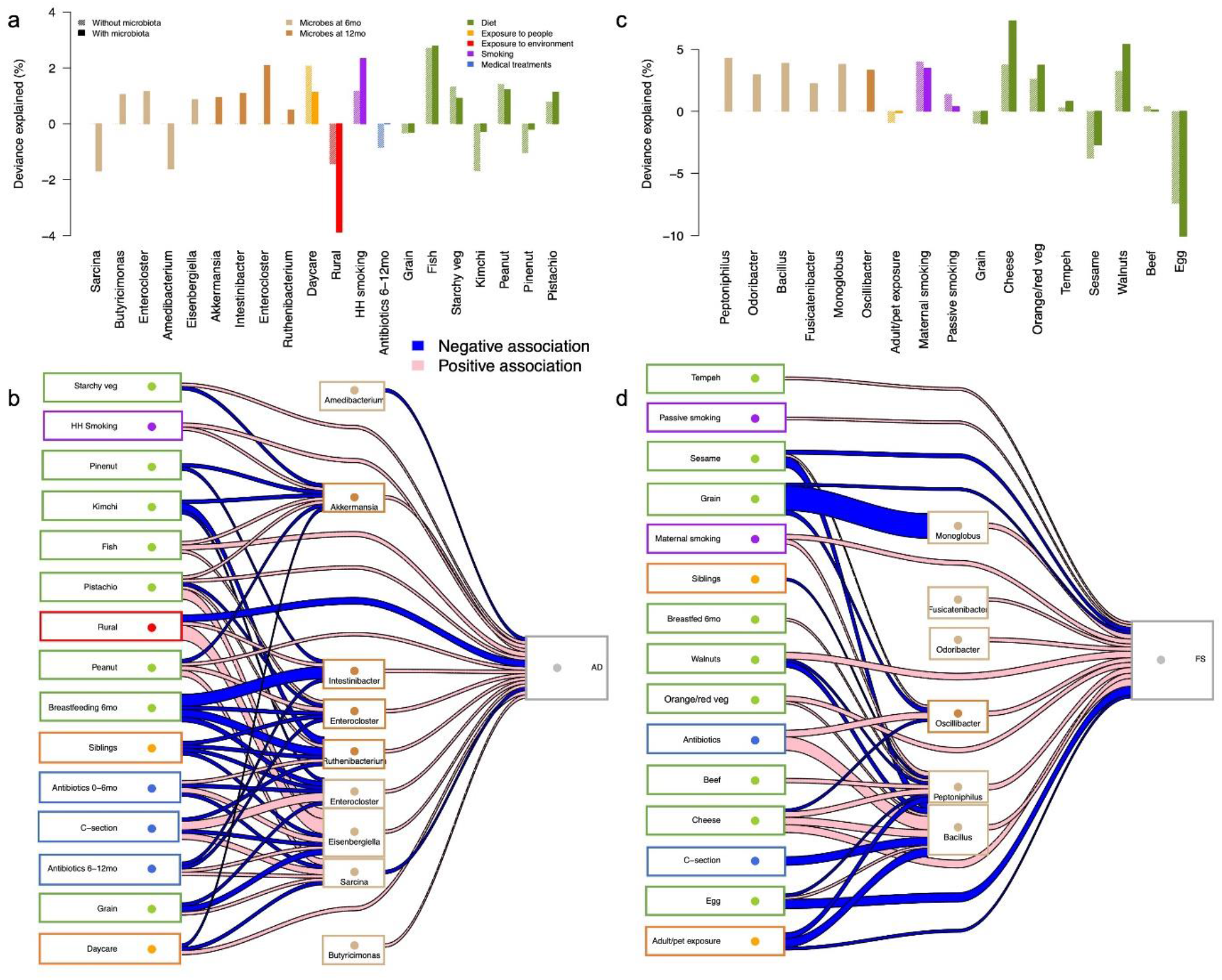
Predictors of atopic dermatitis (a-b) and food sensitization (c-d) based on logistic regression models with and without microbiota. In panels a and c, negative deviance indicates a negative association. Panels b and d show the direct and microbiota-mediated associations between exposures and health outcomes at 1 year. The width of the connecting lines indicates strength of the association (deviance explained).

Interestingly, our analysis revealed that rural living had a protective direct effect on atopic dermatitis, but also a predisposing microbiota-mediated effect through increased relative abundances of *Intestinibacter* and *Eisenbergiella* (**Fig. 5b**). The effects of daycare and antibiotics were microbiota mediated, evidenced by the decrease in effect size when microbiota was included in the model. Attending daycare was associated with reduced relative abundance of *Akkermansia*, and antibiotics reduced the relative abundances of *Intestinibacter* and *Enterocloster* (**Fig. 5b**).

In terms of diet, atopic dermatitis was positively associated with the frequent infant consumption of starchy vegetables, fish, peanuts and pistachios, and negatively with the consumption of grain, pine nuts, and kimchi (**Fig. 5a**). Many foods had both direct and microbiota-mediated effects on atopic dermatitis risk, with the direct and microbiota-mediated effects partly countering each other (**Fig. 5b**). Starchy vegetables and peanuts had a direct predisposing association, but they were also associated with reduced abundances of atopic dermatitis-associated microbes, such as *Akkermansia*. In contrast, the protective effects of kimchi and pine nuts were almost fully explained by their negative impact on *Akkermansia, Intestinibacter, Ruthenibacterium*, and *Enterocloster*. Breastfeeding, siblings, birth mode and antibiotics in the first 6 months did not have a direct association with atopic dermatitis but appeared to modulate risk through their effects on the microbiota.

The AUC for the food allergen sensitization model without gut microbiota was 0.88 (95% CI 0.82-0.94) and adding the gut microbiota increased the AUC to 0.98 (95% CI 0.97-1.00) (**Supplementary Fig 3**). Gut microbiota composition at 6 months emerged as the strongest predictor of food allergen sensitization at 12 months of age, and it appeared to both mediate the effects of several dietary components and exposures, and act as an important confounder for the effects of others (**Fig. 5c**). Food sensitization was primarily associated with the 6-month gut microbiota composition, the predictive microbes being members of Clostridia except for *Bacillus* (Bacilli, *Bacillaceae*) and *Odoribacter* (Bacteroidia, *Odoribacteraceae*). Regarding exposures, having more than 2 adults in the household, or an essential worker, or a pet (adult/pet exposure) had a protective association with food sensitization that was mostly mediated through decreased relative abundances of *Bacillus* and *Peptoniphilus* (**Fig. 5d**). Maternal smoking had a direct influence on food sensitization and an influence mediated by *Peptoniphilus*, while passive smoking only had a direct predisposing effect. In addition, having siblings and C-section birth were associated with reduced relative abundances of food sensitization-associated microbes, while antibiotics in the first 6 months had the opposite effect.

Of the dietary components, tempeh had a direct positive association and egg had a direct negative association with food sensitization (**Fig. 5c-d**). Cheese, orange/red vegetables, and walnuts had a direct positive association, as well as microbiota-mediated effects that were partly protective. The predisposing effect of beef was fully explained by its positive impact on *Peptoniphilus*. Grains and sesame had a direct negative association with food sensitization as well as microbiota-mediated protective effects through reduced relative abundances of *Monoglobus, Oscillibacter*, and *Bacillus*.

## Discussion

In this study, we establish that social isolation due to the COVID-19 pandemic altered the early life assembly of the gut microbiota in infants, which was associated with risk of atopic diseases. We discovered that the primary factors that impact early-life microbiome assembly include specific dietary factors and exposures to other humans.

Furthermore, we established that known epidemiological links between breast feeding, siblings, antibiotics, and diet with risk of atopy are partly mediated via the microbiome. Overall, our findings provide direct support of the ‘metacommunity hypothesis’ of allergic disease whereby lifestyle factors affecting ecological processes that shape the human gut metacommunity at local and wider scales predispose humans to immunological deficits that contribute to allergy risk.

Neonatal gut microbiota is shaped by maternal gut microbes obtained at birth, but the relative contributions of the initial vertical colonization and later horizontal exposures to microbiota maturation in infancy has not previously been well established (36,37). Despite the strict social distancing measures, which were largely complied with in Ireland, the overall gut microbiota composition and development followed closely the expected natural pattern, indicating that the most important source of gut microbes in infants is the mother and other family members (38). During the first months of life, the infant microbiota is dominated by strains from *Actinobacteria* and *Bacteroidia*, which are maternally derived in vaginally born neonates (36). In contrast, maternal strains of *Clostridia* were never observed in neonates suggesting that *Clostridia* are transmitted from environmental sources, perhaps related to lack of persistence previously described for sporulating microbes (39). This supports our findings that environmentally derived taxa, mainly *Clostridia*, are reduced in socially isolated infants, while levels of maternally derived taxa are consistent with historical cohorts. Some species from the *Clostridia* family are well recognized as important immunomodulatory microbes, and altered *Clostridia* levels were previously observed early in life in allergic children (40–43). Our results show that some members of Clostridia that were reduced in the CORAL cohort, were associated with increased risk of atopic outcomes, suggesting potentially protective effects of delayed colonization. This highlights the need to better define the optimal timings for microbiota maturation during infancy.

Higher *Bifidobacterium* levels at 12 months might be related to a reduction in antibiotic use and a high breastfeeding rate in this cohort. Antibiotic use in infancy and later childhood causes a dramatic, long-term reduction in bifidobacteria (44,45). The relative importance of undisturbed gut microbiota development in the first year of life versus delayed *Clostridia* colonization on the long-term health of infants is an important remaining question with implications for infant care practices and decisions, such as entry to daycare and early social contacts. There is likely an optimal age when the benefits of increasing social contacts begin to outweigh the risks of infection and consequent antibiotic exposure. The effect may also vary between outcomes: early-life antibiotic use was associated with a microbiota-mediated risk of obesity in later childhood (45), which is perhaps reduced in the socially distancing cohort, while the risk of immune-related diseases may be increased.

Following acquisition of potential colonizers, our results suggest that the impact of diet on infant gut microbiota development clearly outweighs the effects of social contacts and environment. During breastfeeding, human milk oligosaccharides support specialized microbes, such as bifidobacteria, that protect against infection and support immune development via multiple mechanisms including secretion of metabolites (e.g. SCFAs, indole-3-lactic acid) (46). The introduction of complementary foods leads to a rapid expansion in microbiota diversity and a shift to taxa that can utilize dietary fibers instead of human milk oligosaccharides, accompanied by changes in production of known immunomodulatory microbial metabolites, such as butyrate (47–49). In the CORAL cohort, plant based dietary sources (including legumes, tree nuts and sesame seeds) had the most significant effects on microbiota composition at 6 and 12 months. Of note, the early introduction of potentially allergenic plants foods such as peanut and tree nuts is suggested to protect against food allergy development (50,51). Our study suggests that these foods are utilized by the early life microbiota, which may play a role in their allergy protective effects complementing mechanisms associated with direct sampling by the immune system. It was remarkable to what degree our findings aligned with what is currently considered healthy eating, and foods that support healthy host-microbe interactions (52). Specifically, plant-based foods in general had similar effects as breastfeeding on microbiota composition, which was not observed for animal-based foods within this age group, potentially due to fibers present within the plant-based foods (52,53). This supports current practice to encourage plant based dietary sources during weaning.

The effect sizes of dietary components and other measured exposures were greater than that expected from previous studies (54,55), perhaps due to the reduced overall complexity of exposures contributing to the CORAL infant’s microbiota thereby allowing more accurate identification of effects on specific taxa. In addition, the greatly reduced use of antibiotics in this cohort removed an important confounding factor to gut microbiota development.

Pets and the presence of essential workers who could leave the family home increased the relative abundance of spore-forming, environmentally transmitted bacteria. House type and living in a rural environment also contributed to microbiota composition at 12 months of age. While exposure to biodiverse environments is important to support assembly and maturation of a diverse microbiome, human-adapted symbionts might only be acquired from contact with other humans. Indeed, while the effect of siblings on allergic disease risk were gut microbiota mediated, the protective effect of living in a rural environment was not, indicating that exposure to green environments exerts its protective effects through other mechanisms, but has limited benefits in terms of gut microbiota development.

Social distancing measures designed to limit spread of SARS-CoV-2 were undoubtedly instrumental in preventing excessive morbidity and mortality in Ireland prior to the availability of vaccines. However, it is important to understand more fully how these practices affected the microbiome, and then, in response, to develop public measures and practices that can, if appropriate, increase exposure to beneficial microbes and simultaneously reduce risk of transmission of infectious agents. This cohort of infants born during the pandemic also allowed us to assess the impact of dietary, environmental, and social exposures on development of early life gut metacommunity processes over the first year of life, and their association with immune-mediated disorders such as atopic dermatitis and food allergen sensitization. Whether these differences will be short lived, with rapid recovery post-lockdown, or whether there will be longer term effects remains to be seen.

## Methods

### Study Outline

The CORAL (Impact of **Cor**ona Virus Pandemic on **Al**lergic and Autoimmune Dysregulation in Infants Born During Lockdown) Study is a longitudinal prospective observational study of allergy, immune function and neurodevelopment in a population of term Irish infants born during the first 3 months of the COVID-19 pandemic, March–May 2020. A total of 3773 infants were born in the two participating major maternity hospitals in Dublin from March to May 2020. Invitations were sent to the families of 3065 term babies who were eligible for inclusion. Exclusion criteria were pre-birth PCR-proven SARS-CoV-2 infection in a parent or co-dwelling person, intravenous antibiotics in the neonatal period, multiple births or major congenital anomaly. A total of 351 infants were recruited postnatally to the CORAL Study and mothers did not need to be first-time parents. Babies had a clinical review along with lateral flow COVID-19 antibody screening at 6 and 12 months of age. Detailed epidemiological information was collected at recruitment and at each 6-month review, with a dietary questionnaire performed at 12 months of age.

At recruitment and at each 6-month review, families competed an online questionairre which collected evolving epidemiological information under three main categories. Firstly, the infant’s home environment including household number, presence of siblings, urban or rural location, type of dwelling, childcare outside the home, household pets, and maternal and household smoking status. Secondly, infant health and healthcare utilisation including infant weaning, infant illnesses, antibiotic use, SARS-CoV-2 contacts, testing and positivity, infant hospitalisation, regular medication use and vaccination status. Lastly information was collected regarding symptoms and signs of allergic conditions including suspected adverse food reactions, atopic dermatitis and allergic airways disease.

At 12 month clinical review infants were assessed for signs of atopic dermatitis and severity scoring was completed if present. Skin prick testing was completed to the 3 most common food allergens (peanut, egg and milk) and aeroallergens (house dust mite, grass pollen and cat dander) in all infants.

The food frequency questionnaire measured frequency of intake of baby foods, fruits, vegetables, fermented foods, pulses/legumes, meat, poultry, grains, dairy, eggs, fish and seafoods, peanuts, treenuts and seeds, soy, sugary and fatty foods. We also equired about intake of multivatmins, prebiotics and probiotics. Frequencies included: Avoid, Never, Given and then stopped, Rarely: less than once a month, at least once a month, at least once a week, once a day, more than once a day.

This study involves human participants and ethical permission was granted by the National COVID-19 Ethics Committee (20-NREC-COV-067). Parent/guardian gave informed consent to participate in the study before taking part. Ireland spent the majority of period March 2020–April 2021 in maximal (level 5; no home visitors, stay at home except for essential purposes, exercise within 2–5 km of home, essential retail only, unable to sit in restaurants/cafes, work from home unless essential worker) lockdown and so babies spent most of the first year of their lives in social isolation.

### Microbiota sampling and sequencing

Fecal samples were collected at home using a stool sampling kit with preservative (Norgen Biotek), at 6 and 12 months of age. Stabilized samples were returned via post and stored at −80 C until analysis. Microbial DNA was isolated using the DNeasy PowerSoil Pro Kit (Qiagen) and MiSeq sequencing of the v3 16S rRNA generated approximately 20,000 reads per sample (University of Minnesota Genomics Center).

### Bioinformatic processing

The forward and reverse reads were merged, and the merged reads were quality and chimera filtered using vsearch version 2.14.2 (56). Unique reads occurring only once in the total dataset were removed. Each read was annotated against the RDP 11.5 database using the sintax command in vsearch with 90 % confidence cut-off. The annotated reads were summarized at different taxonomic levels.

### Statistical analysis

The statistical analyses were conducted in R. Principal coordinates analysis (function capscale in R package vegan) using Pearson correlation distances was used to depict the overall gut microbiota composition and its determinants. Functions envfit and adonis in package vegan was used to assess the importance of the background variables on gut microbiota composition. Heat maps were created using the heatmap.2 function in package gplots. Negative binomial models (function glm.nb in package MASS) with the sample-specific read count as an offset were used to identify the factors associated with individual bacterial taxa (57). Logistic regression was used to assess the effects of gut microbiota and diet on the risk of atopic dermatitis and food sensitization. Area under the curve (AUC) was used to assess the predictive ability of the logistic regression models, using the function roc in package pROC (58).

## Data Availability

All data produced in the present study are available upon reasonable request to the authors

## Ethical approval

National Research Ethics Committee for COVID-19-related Research (NREC COVID-19) Approval number 20-NREC-COV-067.

