## Supplemental Fig. for "Gut microbiome predicts atopic diseases in an infant cohort with reduced bacterial exposure due to social distancing"

**Supplementary Fig. 1.** Bacterial community composition in relation to age and birth mode. a) Principal coordinates analysis of the genus-level microbiota using Pearson correlation distances with number of observed genera depicted as interpolated background color and standard deviations of group centroids shown as colored ellipses. b) Average relative abundances of the most abundant bacterial families. V= Vaginal birth, VABX= Vaginal birth with intrapartum antibiotics, CS = C-section birth.

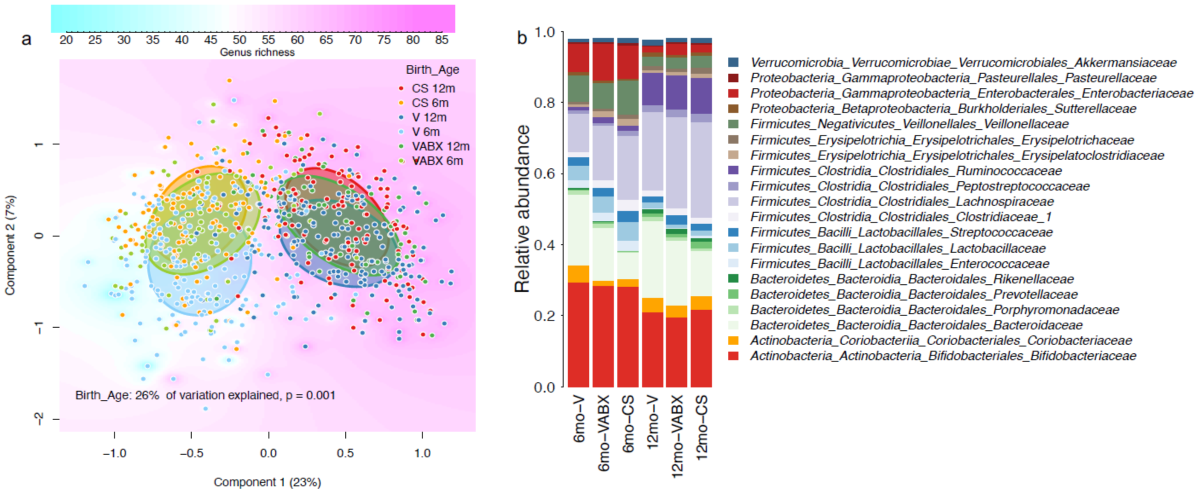

**Supplementary Fig. 2.** ROC curve analysis of the atopic dermatitis prediction model with and without gut microbiota information.

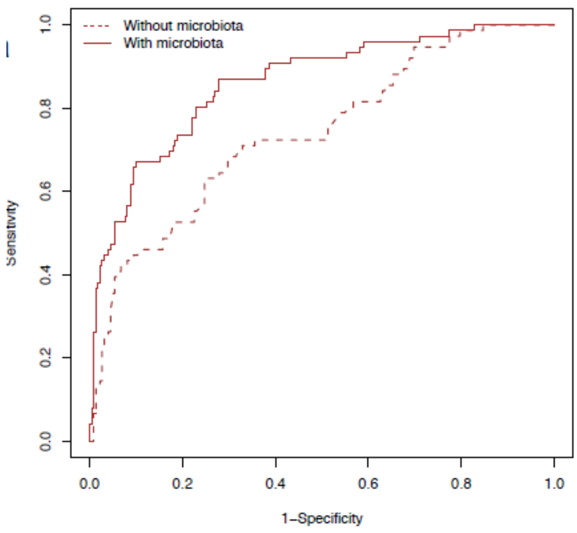

**Supplementary Fig. 3.** ROC curve analysis of the food sensitization prediction model with and without gut microbiota information.

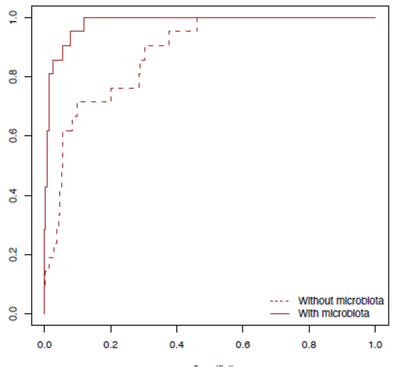
